# Mutant IDH Inhibitors Induce Lineage Differentiation in IDH-mutant Oligodendroglioma

**DOI:** 10.1101/2021.11.16.21266364

**Authors:** Avishay Spitzer, Simon Gritsch, Hannah R. Weisman, L. Nicolas Gonzalez Castro, Masashi Nomura, Nicholas Druck, Rony Chanoch-Myers, Christine K. Lee, Hiroaki Nagashima, Julie J. Miller, Isabel Arrillaga-Romany, David N. Louis, Hiroaki Wakimoto, Will Pisano, Patrick Y. Wen, Keith L. Ligon, Daniel P. Cahill, Mario L. Suvà, Itay Tirosh

## Abstract

Recent data showed promising signs of objective tumor responses in subsets of patients with low grade glioma treated with inhibitors of mutant IDH (IDHi) (1). However, the molecular and cellular underpinnings of such responses are not known. Here, we profiled 6,039 transcriptomes by single-cell or single-nucleus RNA-sequencing isolated from three IDH-mutant oligodendroglioma patients with clinical response to IDHi. Importantly, the tissues were sampled on-drug, four weeks from treatment initiation and our dataset includes a matched pre- and on-treatment sample pair. We integrate our findings with analysis of 8,241 transcriptomes from seven untreated samples, 134 bulk samples from the TCGA and experimental models.(2,3) We find that IDHi treatment induces a robust differentiation towards glial lineages, accompanied by a depletion of stem-like cells and a reduction of cell proliferation. Our study provides the first evidence in patients of the differentiating potential of IDHi on the cellular hierarchies that drive oligodendrogliomas.

## Main text

Hotspot mutations at *IDH1*^*R132*^ and *IDH2*^*R172*^ have been identified in a diverse spectrum of human malignancies, including diffuse gliomas, acute myeloid leukemia (AML), myelodysplastic syndrome (MSD), chondrosarcoma and intrahepatic cholangiocarcinoma.(4–8) These mutations confer a neomorphic enzymatic activity(9), leading to changes in cellular metabolism(10–13) and over-accumulation of the oncometabolite R-2-hydroxyglutarate (2HG).(14) 2HG accumulation has been shown to promote tumorigenesis by competitively inhibiting α-ketoglutarate-dependent dioxygenases, such as histone lysine demethylases and DNA demethylases(15,16), resulting in histone and DNA hypermethylation, thereby altering the cellular epigenetic states.(17–20)

The oncogenicity of mutant IDH, together with its specificity to particular cancer types and ubiquitous expression across malignant cells in these tumors, make it an attractive therapeutic target.(21) Several small-molecule inhibitors of mutant IDH (IDHi) have been developed and are currently undergoing pre-clinical and clinical assessment. While these drugs have shown significant efficacy in IDH-mutant AML,(22–26) preliminary results from phase 1 studies in patients with progressive high-grade gliomas have shown scant signs of activity, and there is a lack of cytotoxic effect in *in vitro* cell culture models, which are typically representative of high-grade disease.(13,27–29) However, recent data from phase 1 studies of the IDHi ivosidenib (AG-120) and vorasidenib (AG-881) (NCT03343197) and BAY1436032 (NCT02746081) showed promising signs of objective tumor responses in a small subset of patients with low-grade gliomas.(1,28) A phase 3 study of vorasidenib in patients with residual or recurrent IDH mutant grade 2 gliomas is currently underway (INDIGO trial, NCT04164901). Yet, the molecular and cellular bases of the response to IDHi in tumors remains uncharacterized.

To examine the cellular and molecular underpinnings of responses to IDHi, we comprehensively analyzed four oligodendroglioma samples (with confirmed *IDH1*^*R132H*^ mutation and chromosome 1p/19q co-deletion) isolated from three patients treated at our institutions who had evidence of clinical response to IDHi (see **Table S1** for clinical information, and **Fig. 1A, B** for study overview and timelines)(1). Patient MGH170 showed partial response as measured by RANO criteria after 10 months of treatment with IDHi and patient MGH229 showed stable disease without progression after 11 months of IDHi treatment. Under existing sampling protocols, tumor tissue was obtained from MGH170 and MGH229 on-treatment 4 weeks after initiation of IDHi. These tumor samples were profiled by single-cell RNA-sequencing (scRNAseq) using the full-length SMART-Seq2 protocol.(30) Overall, 1,153 cells passed our stringent quality controls, with 4,333 genes detected per cell on average, underscoring the high-quality of our dataset. We further profiled a matched pair of frozen samples (pre- and on-treatment) from a third oligodendroglioma patient (BWH445) who showed clinical response to IDHi treatment by single-nucleus RNA-sequencing (snRNAseq, 10x genomics). Patient BWH445 showed stable disease without progression after 30 months from initiation of IDHi treatment (**Fig. 1B, Table S1**). The on-treatment sample from BWH445 was obtained 4 weeks after initiation of IDHi, as in the former two patients. Overall, 8,793 nuclei passed quality control with 2,816 genes detected on average.

**Figure 1.**
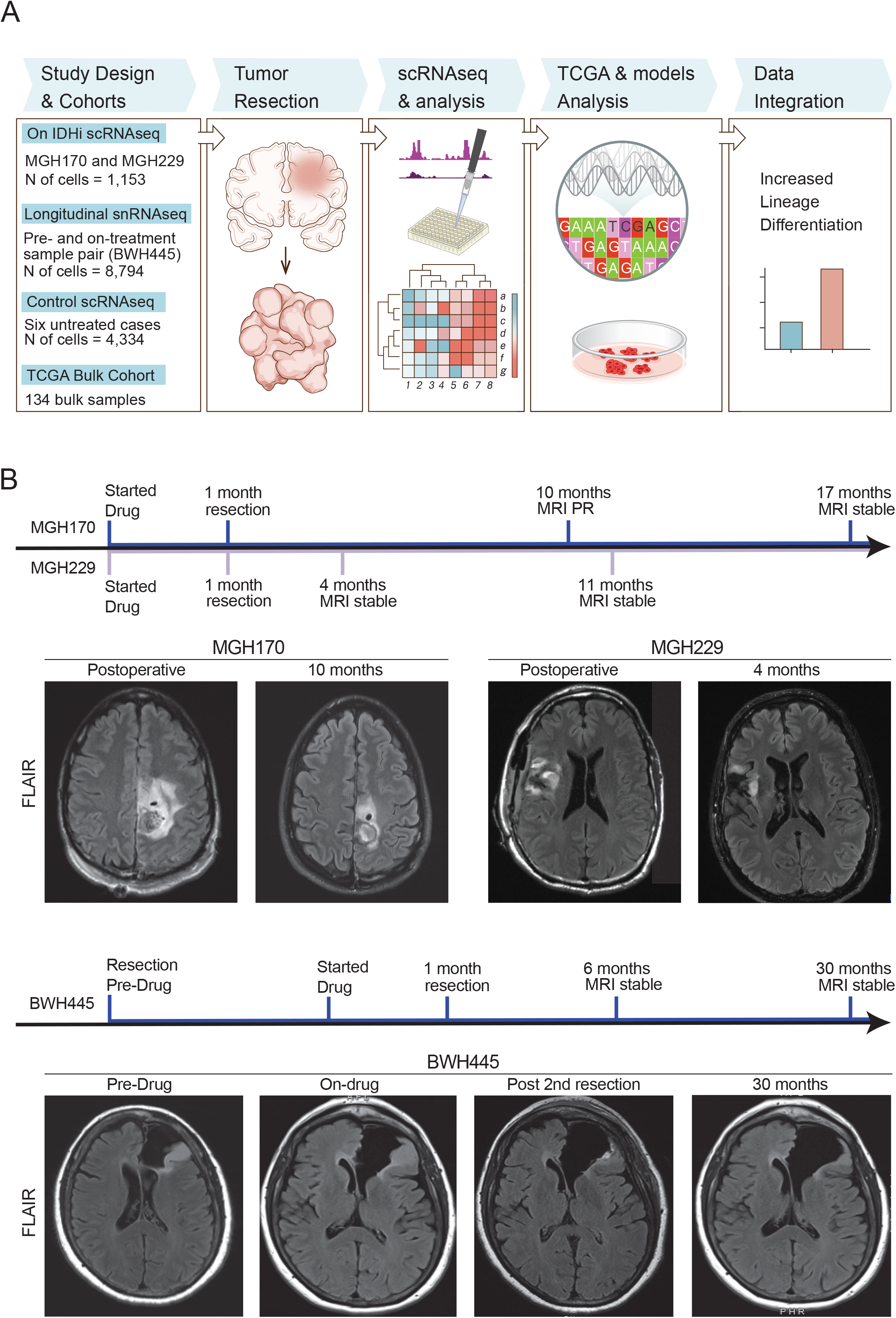
Study workflow and clinical timeline. (**A**) Scheme describing the workflow of this study. (**B**) Top panel shows an overview of the clinical history of MGH170 and MGH229 after initiation of IDHi treatment as well as brain MRIs of the two patients treated with IDHi. Patient MGH170 achieved partial response (PR) as measured by RANO criteria after 10 months of treatment with IDHi while patient MGH229 showed stable disease without progression after 11 months of treatment with IDHi. Bottom panel shows the clinical history of BWH445 before and after initiation of IDHi therapy as well as representative brain MRIs of this patient. Patient BWH445 showed stable disease without progression after 30 months of treatment with IDHi.

The two IDHi-treated glioma samples profiled using scRNAseq were combined with 4,334 single-cell transcriptomes that we previously obtained from six treatment-naïve grade 2 oligodendrogliomas (**Fig. 2A**)(2,3) whereas the matched pair was analyzed separately from this cohort by comparing between the two timepoints (**Fig. 2B**). As we previously described, malignant glioma cells were clearly distinguishable from the non-malignant cells (immune cells and glial cells) by inferred copy number aberrations (CNA), including the oligodendroglioma defining co-deletion of chromosome arms 1p/19q, and by expression of marker genes (**Fig. 2C, S1A-B, Table S2**).(2)

**Figure 2.**
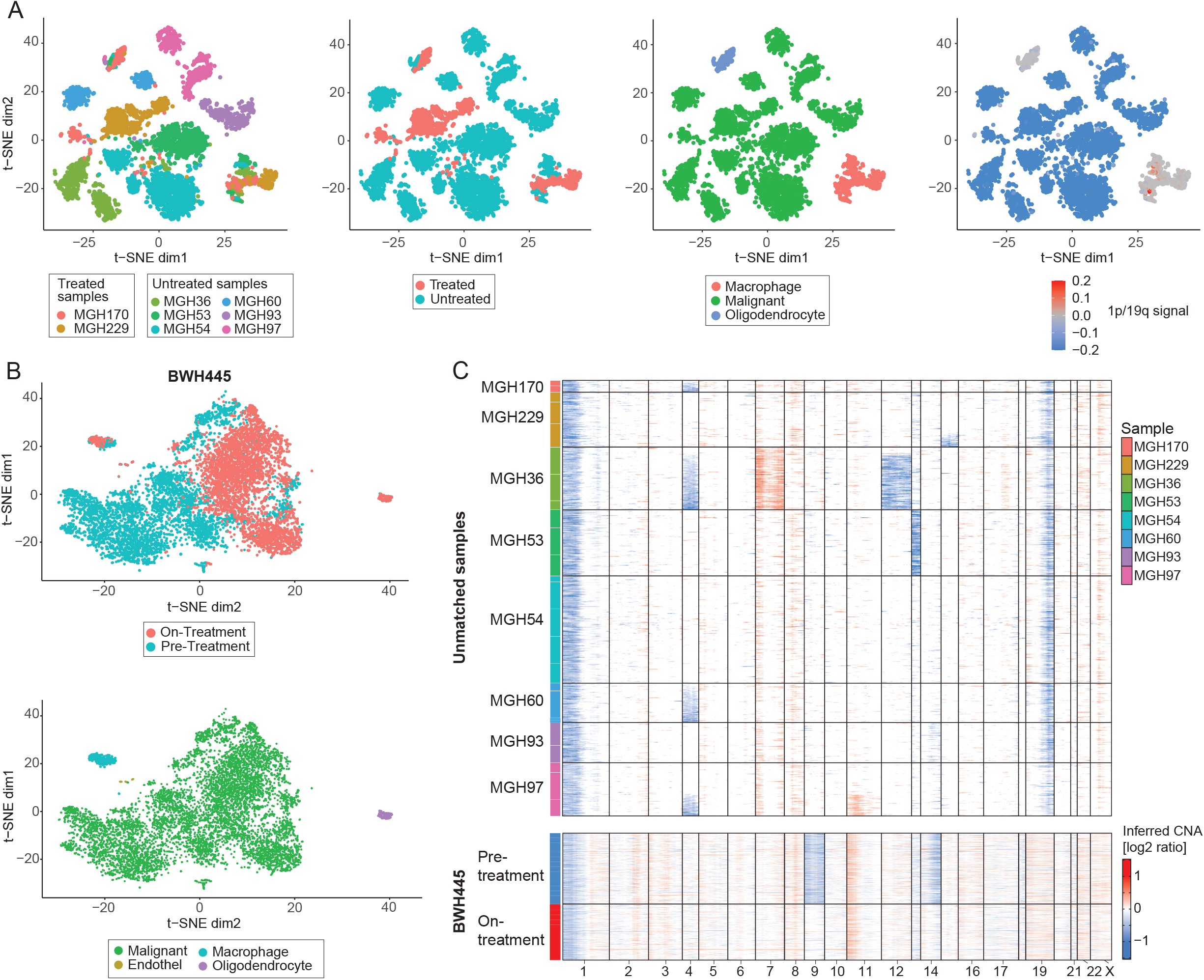
Profiling of patients with clinical response to IDHi. **(A)** t-distributed stochastic neighbor embedding (t-SNE) plots of the *unmatched samples* showing 5,487 single cell expression profiles. Each of the four plots shows the same cells and coordinates, but colored in a different way as labeled below the figure, with colors corresponding to (from left to right): sample identity, treatment status, cell type and co-deletion of chromosomes 1p/19. (**B**) t-distributed stochastic neighbor embedding (t-SNE) plots showing 8,219 single cell expression profiles of the *matched on- and pre-treatment samples (BWH445)*. Each of the two plots shows the same cells and coordinates, but colored in a different way as labeled below the figure, with colors corresponding to (from left to right) treatment status and cell type. (**C**) Copy-number aberrations inferred from single-cell expression data of cells classified as malignant. Rows represent cells and columns represent chromosomal location. The expected co-deletion of chromosome arms 1p and 19q is clearly seen in the unmatched samples as well as some other aberrations that tend to be subclonal (see **Fig. S1A** for CNA profile of the non-malignant cells). For the matched samples (BWH445) only the 1p deletion could be inferred from the single-cell expression data (co-deletion was validated using array comparative genomic hybridization, see panel **S1F**).

Comparison between IDHi-treated and untreated samples (non-paired samples) using gene-set enrichment analysis (GSEA, see *Methods*) with a strict significance threshold (adjusted p-value < 0.05) identified 230 enriched gene-sets (**Fig. 3A, Table S3**). These included gene-sets defining the three central components of the cellular hierarchy that we previously proposed for IDH mutant glioma: proliferating cells resembling neural progenitor cells (NPC-like) and cells differentiated towards the astrocytic (AC-like) and oligodendrocytic (OC-like) lineages.(2) The most highly enriched gene-sets in IDHi-treated samples were those of the AC-like states previously defined in glioma as well as gene-sets of normal astrocytic lineage differentiation.(2,3,31–37) Conversely, the most highly enriched gene-sets in the untreated samples were of the OC-like state in glioma. When examining the differentially expressed genes (DEGs) between treated and untreated samples we cannot discern their statistical significance due to the limited sample size (cells from the same patient cannot be considered statistically independent and hence we compare 2 treated with 6 untreated pseudo-bulk profiles). However, when focusing on genes with the highest fold changes, AC-like genes account for a large fraction of the upregulated genes while OC-like and NPC-like genes account for a large fraction of the downregulated genes (**Fig. 3B**). Specifically, 457 genes defining the AC-like, OC-like and NPC-like states together account for 53% of the 102 most DEGs (with an average of 4-fold expression difference), compared with the 4.7% expected by chance (p=4.92^-15^ by hypergeometric test). Comparison between the pre- and on-treatment samples of BWH445 using GSEA revealed the same pattern of enrichment with astrocytic gene-sets upon IDH inhibition (**Fig. 3C**). Moreover, when combining the DEG lists of the unmatched and matched cohorts (see *Methods*) we found that this combined list was enriched (70/152) with core genes of the AC-like (*AQP4, MT1X, CST3, NTRK2*), OC-like (*DLL3, SOX8*) and NPC-like (*DCX, ASCL1, HES6, ELAVL4*) expression programs (**Fig. 3D**). This striking enrichment suggests that *in vivo* IDH inhibition in glioma patients primarily affects these neurodevelopmental programs.

**Figure 3.**
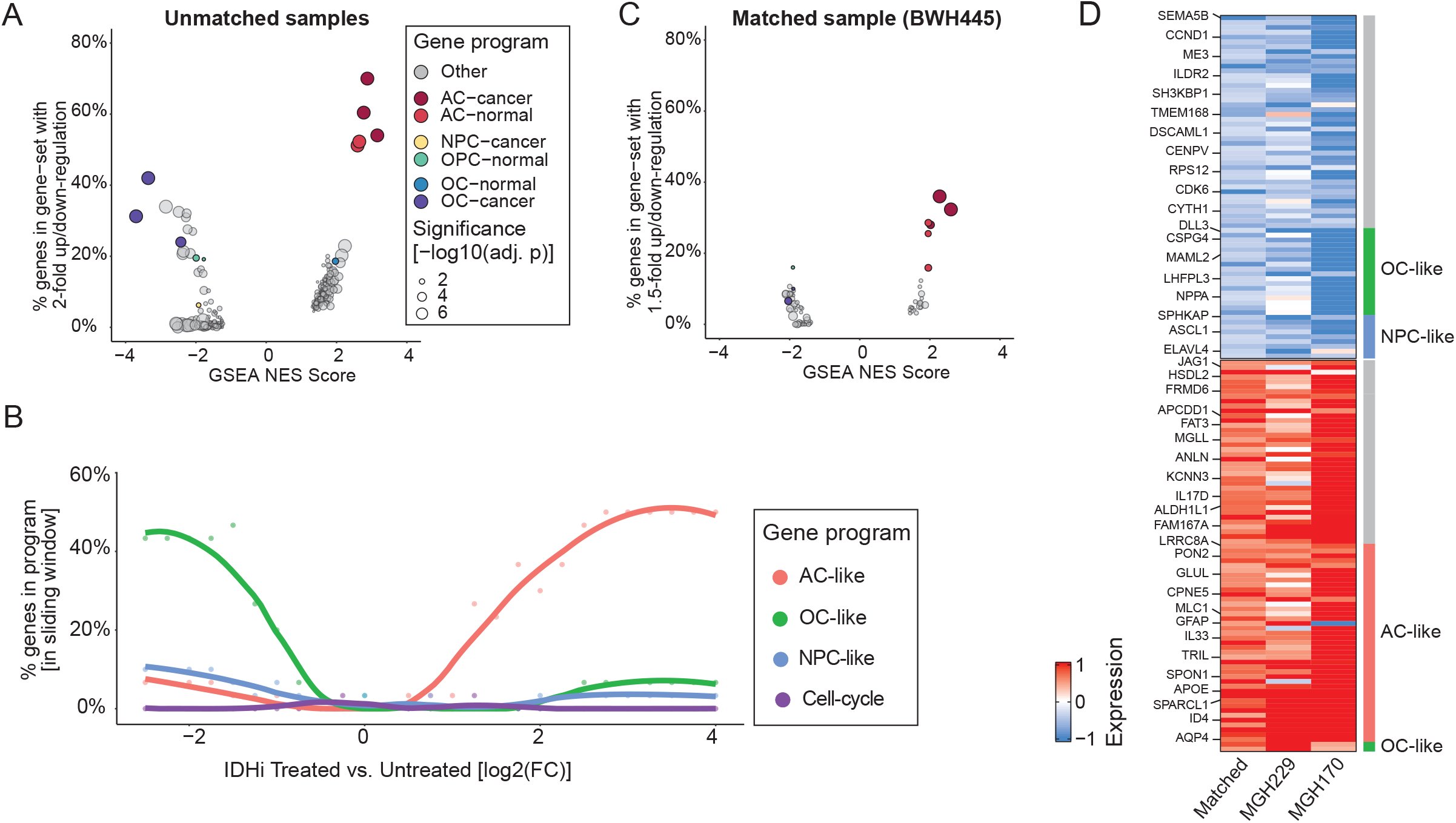
Gene-set enrichment analysis. (**A**) Comparison of IDHi-treated and untreated samples using GSEA. Each dot represents a gene-set that passed the statistical significance threshold (adjusted p-value < 0.05), dot size represents the extent of statistical significance and dot color indicates whether the gene-set belongs to any of the glioma hierarchy/neural development gene-sets. X-axis shows the GSEA Normalized Enrichment Score (NES), Y-axis shows the fraction of genes in each gene-set with an absolute log2-ratio greater than 1 (e.g. genes in the extreme ends of the ranked list used for GSEA computation with more than a twofold change). (**B**) Enrichment of the ranked list used for GSEA (unmatched cohort). Dots represent the percentage of genes, in a sliding window of 30 genes (by log2-ratio values), that overlap each of the three expression programs and the cell cycle. Trend line was computed using LOESS regression. (**C**) Comparison of matched on- and pre-treatment samples using GSEA (same as panel A). Y-axis shows the fraction of genes in each gene-set with an absolute log2-ratio > log2(1.5) (e.g. 1.5 fold up/down-regulation). (**D**) Common genes of the most highly DEGs (computed separately for the unmatched and matched cohorts). Rows represent samples, columns represent DEGs, which are annotated according to the expression programs with which they are associated. The expression values of the unmatched samples are shown relative to the average bulk expression profile of the untreated samples and those of the matched on-treatment sample are shown relative to the expression profile of its pre-treatment counterpart.

Importantly, these DEGs were not uniformly higher (or lower) in IDHi-treated, compared to untreated, malignant cells. Instead, these genes were highly expressed in specific subsets of cells (AC-like, OC-like and NPC-like) that were all found in both conditions (**Fig. 4A-B**). This raises the possibility that IDHi treatment results in the induction of detectable perturbations in these cellular hierarchies. Therefore, we next examined the effect of IDHi treatment on proportions of these states, as well as of an intermediate “undifferentiated” state and of cycling cells (**Fig. S1C-D**). The IDHi-treated samples had the largest proportion of AC-like cells (p=0.02, one-sided t-test), the smallest proportion of cells classified as either Undifferentiated or NPC-like (p=0.002, one-sided t-test) and the smallest proportion of cycling cells (p=0.01, one-sided t-test). This pattern was largely evident in any pairwise comparison between an IDHi-treated and untreated samples (**Fig. S3A**). A potential caveat in our comparisons of IDHi-treated to untreated patient samples, is that the former patient group has progressed on standard treatment before being treated by IDHi, while the latter patient group was profiled prior to progression. However, we note that progression in IDH-mutant glioma is linked to a decrease in glial differentiation, as we demonstrated previously (3) and as inferred from analysis of TCGA samples (see **Fig. S5A**). Thus, IDHi-treated samples would be expected to show fewer AC-like cells (than in untreated samples), suggesting that the observed increase in AC-like cells is directly linked to their treatment by IDHi. This conclusion is further supported by comparison of the matched pre- and on-treatment BWH445 samples, which revealed a 2-fold increase in the proportions of both AC-like and OC-like cells and a 4-fold decrease in the proportion of cycling cells (**Fig. 4C-D, S2B-D**).

**Figure 4.**
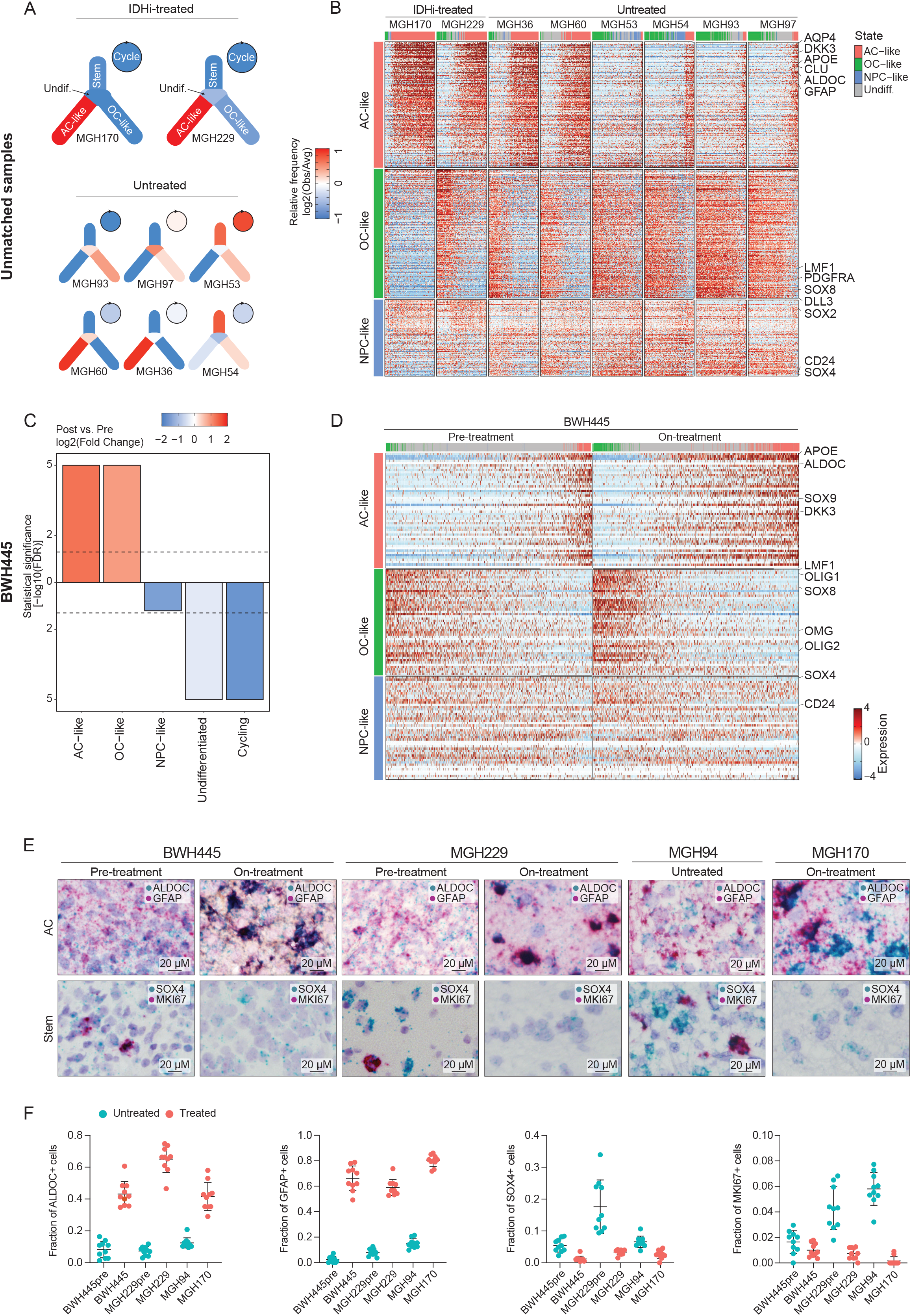
IDHi treatment is associated with AC differentiation. (**A**) Scheme depicting a model of cellular hierarchy in which colors represents the relative frequency of each cellular state in each tumor sample of the unmatched cohort. (**B**) Relative expression of genes associated with the AC-like, OC-like and NPC-like programs across the unmatched samples. Cells are ordered by AC-like score minus OC-like score; genes are ordered by expression log2-ratio. For visualization, each tumor was randomly down-sampled to 130 cells. (**C**) Comparison of the fraction of cells assigned to each state between the on- and pre-treatment matched samples. Bar values represent the statistical significance of each pairwise comparison, defined as -log10 of a p-value calculated by hypergeometric test and corrected for multiple testing using the Benjamini-Hochberg method; bar direction (up or down) is defined by an increase or decrease, respectively, of the relevant state fraction in the on-treatment sample. Bar colors represent the relative change in state fractions (computed as 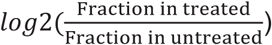. (**D**) Relative expression of genes associated with the AC-like, OC-like and NPC-like programs in the matched on- and pre-treatment samples (same as panel C, no down-sampling in this case). (**E**) *In situ* RNA hybridization of tumors BWH445 (pre-treatment and on-treatment), MGH229 (pre-treatment and on-treatment), MGH94 (untreated) and MGH170 (on-treatment) for AC-like (*GFAP, ALDOC*), Stem-like (*SOX4*), and proliferation (Ki67) markers. (**F**) The fraction of cells staining positive for *GFAP, ALDOC, SOX4* and Ki67 based on RNA *in situ* hybridization of tumors BWH445 (pre-treatment and on-treatment), MGH229 (pre-treatment and on-treatment), MGH94 (untreated) and MGH170 (on-treatment). Each dot corresponds to one field of view.

We confirmed our observations by RNA *in situ* hybridization with markers of AC-like cells (*GFAP, ALDOC*), NPC-like cells (*SOX4*) and cell proliferation (Ki-67) in the pre- and on-treatment matched samples, in the two IDHi-treated unmatched samples, in a sample taken from patient MGH229 long before initiation of IDHi treatment (MGH229pre) and in six untreated oligodendroglioma samples (including one additional untreated sample - MGH94) (**Fig. 4E-F, S1** *Methods*). Furthermore, when comparing cells classified into the same state, we noticed differences between IDHi-treated and untreated cells that further supported an induction of differentiation by IDHi. Cells classified as AC-like in two IDHi-treated samples (MGH170 and MGH229) were enriched with gene-sets reflecting normal astrocytic development, compared to AC-like cells in untreated samples (**Fig. S3B**). Thus, IDHi may promote both an increased *proportion* of AC-like cells and an increased *degree* of differentiation, such that AC-like differentiation in IDHi-treated samples better recapitulates the normal astrocytic state. Interestingly, a similar pattern was observed for cells classified as OC-like, which displayed enrichment of gene-sets reflecting normal oligodendrocytic development(31–36) in IDHi-treated vs. untreated samples (**Fig. S3C-D**). However, unlike the consistent increase in AC-like differentiation, the effect on OC-like differentiation appears to be variable, with increased proportion of OC-like cells in BWH445, apparently decreased proportion in MGH170 and decreased proportion but increased differentiation in MGH229. Taken together, these results suggest that response to IDHi treatment results in the induction of cellular differentiation. While there are multiple components to such induction of differentiation, the predominant consequence is an increase in the fraction and degree of differentiation of AC-like cells (and more variably of OC-like cells), and a decrease in the proportion of undifferentiated and proliferating cells.

A strength of our study is the analysis of human glioma tissue on-treatment, including an analysis of a matched pre-and on-treatment sample pair. However, this work has two important limitations - a small sample size and lack of a model system - that we describe below and attempt to ameliorate by further analysis.

The limited size of our cohort, along with the unique clinical course of each patient in the cohort (**Table S1**) are inherent limitations of our study. However, profiling of human glioma tissue on-treatment has been a major limitation in trials due to limited indication for reoperation. Additionally, studies of IDHi showed signs of tumor responses only in a small subset of patients, making acquisition of on-treatment tissue from patients with clinical response to IDHi treatment even more challenging. To partially overcome this limitation and increase the confidence in our results, we performed two additional analyses. First, we performed a similar scRNAseq analysis of four samples from three IDH-mutant astrocytoma patients that were treated by IDHi but had no clinical response. Importantly, this analysis did not show an increase in lineage differentiation as observed for the responders (**Fig. S4**). Second, we extended our analysis by leveraging bulk RNA-seq profiles of 134 oligodendrogliomas from the TCGA lower-grade glioma cohort. Using the signatures derived from scRNAseq, we could score each bulk tumor for its overall degree of AC-like differentiation, and compared this to the AC-like differentiation scores of our scRNAseq cohort, when averaging across the cells in each tumor (**Fig. S5A**; *Methods*). Consistent with our previous observations (with 6 untreated samples), we found that the IDHi-treated samples were highly AC-differentiated when compared to 134 untreated TCGA samples (**Fig. S5B**). Specifically, MGH170 (partial clinical response) was the most AC-differentiated among all samples, whereas MGH229 (stable disease) was within the top 3% of AC-differentiated samples (4 out of 134). By comparison, our untreated samples analyzed by scRNAseq spanned the expected spectrum of AC-like differentiation within the TCGA cohort (p=0.54 for difference between groups, Wilcoxon rank sum test).

Our second limitation is the lack of a robust cellular model to validate our findings *in vitro* and to further investigate the underlying mechanisms. Cell lines are notoriously difficult to derive from IDH-mutant 1p/19q co-deleted tumors. We therefore utilized the closest models that we could obtain - gliomasphere cultures derived from three IDH-mutant *non-1p/19q co-deleted* patients. scRNAseq analysis of these models demonstrated that they partially recapitulate the cellular states observed in patient IDH-mutant gliomas (**Fig. S6A-C**, *Methods*). We then compared the expression of IDHi-treated to untreated cells by scRNAseq in each model independently. In all three of these models, IDHi treatment was associated with an increase in the fraction of AC-like and/or OC-like cells and a decrease in the fractions of undifferentiated, NPC-like and cycling cells (**Fig. S6D-F, Table S4**). This effect was considerably smaller than observed in the patient samples, likely reflecting the limitations of such *in vitro* models, which lack 1p/19q co-deletion, lack important micro-environmental components and are biased towards advanced stages of the disease where response to IDHi may be reduced due to additional progressive mutations(13,38,39).

Our observation that inhibiting the activity of a driver mutation induces differentiation is consistent with a model of oncogenesis through a block of differentiation(18,40). Notably, induction of differentiation could provide clinical benefits that may not be properly assessed by measures of response that have been designed for cytotoxic drugs, which could potentially explain some of the controversy around the efficacy of IDHi.(13,27–29,41) Nonetheless, radiographic response to IDHi is limited to a subset of IDH-mutant glioma, highlighting the important need of identifying which patients will respond.

Since response to IDHi may require plasticity along AC-like differentiation states, pre-existing diversity of such states in untreated samples may predict the capacity for further differentiation and hence the response to IDHi. AC differentiation is decreased in gliomas with increased grade(3) (**Fig. S5A**), suggesting that IDHi efficacy might be limited to low-grade gliomas. This is consistent with the increased response rate observed in non-enhancing compared to enhancing IDH-mutant glioma(1), as well as with the stronger effect that we observed in MGH170 (grade 2) compared with MGH229 (grade 3). Given the potential significance of AC differentiation for IDHi response, we utilized the TCGA cohort to examine if particular mutations correlate with AC differentiation. We found that mutations in *NOTCH1* are associated with a low degree of AC differentiation in grade 2 lesions (**Fig. S7A-C, Table S5**), suggesting *NOTCH1* mutation and/or activation may serve as a potential biomarker for stratifying the response to IDHi.

In summary, our study provides a proof-of-concept for targeted differentiation therapy with mutant IDH inhibitors directly in IDH-mutant glioma patients and sheds light upon the underlying transcriptional and epigenetic changes. While differentiation of malignant cells in patients treated with IDHi has been observed in AML(23), this is to our knowledge the first demonstration of such a response in solid tumors in patients. Understanding the cellular response to IDHi treatment will help (i) identify those patients more likely to benefit from it, (ii) assess response and resistance to differentiation therapy in low-grade gliomas and (iii) aid the identification of potential targets for combination strategies.

## Methods

### Acquisition and processing of human glioma samples for single cell RNAseq

Patients at Massachusetts General Hospital (MGH) were consented preoperatively in all cases according to Institutional Review Board DF/HCC 10-417. Fresh tumors were collected at the time of surgery and presence of glioma was confirmed by frozen section. Tumor specimens were mechanically dissociated with a disposable, sterile scalpel and further enzymatically dissociated into single cell suspensions using the enzymatic brain dissociation kit (papain-based) from Miltenyi Biotec as previously reported(2,3,42,43). Tumor cells were blocked in 1% bovine serum albumin in phosphate-buffered saline solution (1% BSA / PBS). Cell suspensions were subsequently stained for flow cytometry for 30 min at 4 °C using antibodies specific for CD45 [REA747]-VioBlue and CD3 [BW264/56]-PE from Miltenyi Biotec. Cells were washed with cold PBS, and then incubated for 15 min in 1.5 mL of 1% BSA / PBS containing 1 uM calcein AM (Life Technologies) and 0.33 uM TO-PRO-3 iodide (Life Technologies). Sorting was performed with the FACS Aria Fusion Special Order System (Becton Dickinson) using 488 nm (calcein AM, 530/30 filter; CD3-PE, 585/42 filter), 640nm (TO-PRO-3, 670/14 filter), and 405 nm (CD45-VioBlue, 450/50 filter) lasers. Standard, strict forward scatter height versus area criteria were used to discriminate doublets and gate only singleton cells. Viable cells were identified by staining positive with calcein AM but negative for TO-PRO-3. We sorted individual, viable, CD45+CD3- and CD45+CD3+ immune, and CD45-non-immune single cells into 96-well plates containing cold TCL buffer (QIAGEN) with 1% beta-mercaptoethanol. Plates were frozen on dry ice immediately after sorting and stored at -80 °C prior to whole transcriptome amplification, library preparation and sequencing.

### RNA in situ hybridization

Paraffin-embedded tissue sections from human tumors from Massachusetts General Hospital were obtained according to Institutional Review Board-approved protocols. Sections were mounted on glass slides and stored at −80°C. Slides were stained using the RNAscope 2.5 HD Duplex Detection Kit (Advanced Cell Diagnostics, Cat. No. 322430). Slides were baked for 1 h at 60°C, deparaffinized and dehydrated with xylene and ethanol. The tissue was pretreated with RNAscope Hydrogen Peroxide (Cat. No. 322335) for 10 min at room temperature and RNAscope Target Retrieval Reagent (Cat. No. 322000) for 15 min at 98°C. RNAscope Protease Plus (Cat. No. 322331) was then applied to the tissue for 30 min at 40°C. Hybridization probes were prepared by diluting the C2 probe (red) into the C1 probe (green). The ratio of C2:C1 was 1:50 for MKI67/SOX4 and 1:60 for GFAP/ALDOC. Advanced Cell Technologies RNAscope Target Probes used included Hs-GFAP-C2 (Cat No. 311801-C2), Hs-ALDOC (Cat No. 407031), Hs-SOX4 (Cat No. 469911), Hs-MKI67-C2 (Cat No. 591771-C2). Probes were added to the tissue and hybridized for 2 h at 40°C. A series of 10 amplification steps were performed using instructions and reagents provided in the RNAscope 2.5 HD Duplex Detection Kit. Tissue was counterstained with Gill’s hematoxylin for 25 s at room temperature followed by mounting with VectaMount mounting media (Vector Laboratories). For RNA *in situ* hybridization quantification, at least 1,000 cells were counted across 10 high power fields in representative areas of the tumors.

### Single-cell RNAseq data processing of human glioma samples

Smart-seq2 whole transcriptome amplification, library construction, and sequencing were performed as previously published(2,3,42,43). We processed sequencing data from raw reads to gene expression matrices as previously described(2). We used bcl2fastq to generate demultiplexed FASTQ files, and aligned the resulting paired-end scRNAseq reads to the human transcriptome (hg19) using Bowtie (v0.12.7)(44). Statistical analysis was done using *R* version 4.0.1. We merged the gene expression levels which were calculated by RSEM(45) for MGH170 and MGH229 with the ones calculated for the reference samples from *Tirosh et al. 2016*(2) (downloaded from https://singlecell.broadinstitute.org/single_cell). Analysis was done mainly using the R package *scandal* which is freely available at https://github.com/dravishays/scandal. Gene expression levels were quantified as 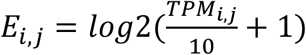 where *TPM*_*i,j*_ refers to transcript-per-million for gene *i* in sample *j*. We divided the *TPM* values by 10 as the size of single-cell libraries is estimated to be in the order of 100,000 transcripts and we therefore would like to avoid inflating the expression levels by counting each transcript ∼10 times. We filtered out cells with fewer than 3000 detected genes yielding 1153 IDHi-treated cells and 4334 untreated cells (**Table S2**). Next, we computed the average expression of each gene *i* as 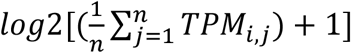 in each tumor sample separately and excluded genes with average expression below 4. We then merged the gene lists from the 8 tumor samples, leaving a set of 10516 genes for downstream analysis. For the cells and genes that passed these quality control procedures we defined relative expression levels by centering the expression levels for each gene across all cells in the dataset as follows: 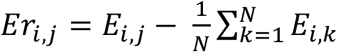 where *N* is the number of cells in the dataset.

### Clustering and identifying presumed normal and malignant cells

We clustered all cells using graph-based clustering (Louvain’s algorithm) of a 2-dimensional UMAP space (R’s UMAP package, distance metric set to “pearson”) computed from the relative expression levels. We identified two small clusters containing cells highly expressing markers of Oligodendrocytes and Macrophages which were annotated accordingly, whereas the rest of the cells were annotated as presumed malignant (**Fig. 1C**).

### CNA analysis

CNAs were estimated as described elsewhere(2,3,37,42,43). Briefly, the algorithm sorts the analyzed genes according to their chromosomal location and applies a moving average with a sliding window of 100 genes within each chromosome to the relative expression values. The scores computed for the cells classified as non-malignant (Oligodendrocytes and Macrophages in this case) define the baseline of normal karyotype and their average CNA values are used to center the values of all cells. Classification of cells as malignant/non-malignant was based on two metrics:

1. CNA signal: reflects the extent of CNAs in each cell, and defined for each cell *j* as 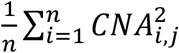 where *n* is the number of genes and.
2. CNA correlation: defined as the Pearson correlation between the CNA profile of each cell and the average CNA profile of all presumed malignant cells from the corresponding tumor sample.

Cells were classified as malignant if CNA signal was above 0.025 and CNA correlation was above 0.25. Cells that failed to pass the two thresholds were marked as “unresolved” and excluded from downstream analysis.

### Gene-sets associated with malignant cellular states

For scoring samples (including TCGA samples) we used gene-sets curated from *Tirosh et al. 2016*(2) denoted here as the *IDH-O AC-like, OC-like, NPC-like, G1/S* and *G2/M* programs. For functional annotation of gene lists such as in **Fig. 2A** we also included gene-sets curated from *Venteicher et al. 2017*(3) and *Neftel et al. 2019*(37). See the gene-sets in **Table S2**.

### Gene-sets associated with normal brain cell types

We scored the tumor samples for gene-sets representing normal cell types which were curated from published scRNAseq datasets(31–36). Each dataset was converted to CPM units (if not already in TPM units) and log2-transformed. Genes were filtered by *Ea* with a mean expression cutoff of 4. Gene-sets were then defined by differential expression between the clusters that were defined in the respective paper. We selected the top 50 genes by log2 fold-change (FC), with log2FC>=1 and Benjamini-Hochberg adjusted p-value<0.05. See the gene-sets in **Table S2**.

### Assignment of cells to states

Malignant cells were scored for the *IDH-O AC-like, OC-like* and *NPC-like* programs using the function *sigScores* from the R package *scalop* available at https://github.com/jlaffy/scalop. We generated 100 shuffled expression matrices by sampling each time 1000 cells and shuffling the expression values for each gene. We then scored each shuffled matrix for the AC-like, OC-like and NPC-like expression programs, thus yielding 100,000 normally distributed scores for each expression program. These were used as null distributions for cell state classification. For each original cell, we computed a p-value for each of the expression programs with a Z-test (R’s *pnorm* function) using the statistics of the null distributions that we previously generated. We adjusted all p-values for multiple testing using the Benjamini-Hochberg method. Each cell was classified into a specific state if the adjusted p-value computed for that state was smaller than 0.05. Cells that either did not achieve an adjusted p-value < 0.05 for any of the states or achieved an adjusted p-value < 0.05 for multiple states (∼6% of cells) were assigned an “Undifferentiated” state. See **Fig. S1D** for the statistics of state assignment for each sample.

### Cell cycle analysis

Malignant cells were scored for the *IDH-O G1/S* and *G2/M* programs and classified as cycling using the same method described above for cell state classification. See **Fig. S1C** for the statistics of cell cycle assignment for each sample.

### Gene-set enrichment analysis

For each sample *s* we computed a bulk expression profile for each gene *i* as 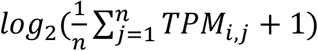 where *n* is the number of cells in sample *s*. We then computed the average *log*_*2*_ expression of each gene across the IDHi-treated and untreated samples and then computed the log_2_-ratio of each gene by subtracting the average log_2_ expression of the untreated samples from that of the treated samples. We then generated the ranked list used for GSEA(46,47) by sorting the log_2_-ratio list. GSEA was computed using the R package *fgsea*(48). We included in this analysis 10264 gene-sets (10185 GO gene-sets, 50 mSigDB hallmark gene-sets, 11 gene-sets reflecting Glioma cellular hierarchy and 18 gene-sets reflecting normal astrocytic, oligodendrocytic and OPC development) which were downloaded from mSigDB or curated as described above.

### Ranked list enrichment analysis (pertaining to Fig. 2A, B -lower panel)

Ranked list genes (see *Gene-set enrichment analysis*) were annotated according to inclusion in AC-like, OC-like and NPC-like gene-sets curated as described above. We computed the fraction of genes associated with a particular state in windows of 30 genes at fixed intervals of 0.25 along the spectrum of log2-ratio values (ranging from -2.5 to 2.5). See **Table S2** that contains the computed fractions.

### Combined DEG analysis of unmatched and matched cohorts

As before-mentioned, the unmatched and matched cohorts were analyzed separately, yielding two lists of DEGs (for the unmatched cohort by comparing the bulk expression profiles of the IDHi-treated samples with those of the untreated samples and for the matched cohort by comparing the expression profiles of the on- and pre-treatment samples). From each of these lists we included in the analysis the 5% most upregulated and 5% most downregulated genes (e.g. 10% of genes) and generated the combined DEG list by intersecting the two lists.

### Permutation test for OPC-OC differentiation (pertaining to Fig. S2E)

Malignant cells were scored for normal OC and OPC programs curated from normal brain datasets as discussed above. We computed the difference between the two scores for each cell and then averaged the score difference per sample. We then computed a sampling distribution of the mean *OC-OPC* score difference by generating 1000 random gene expression matrices as described above. We scored each of these shuffled matrices for the normal OC and OPC expression programs and then counted how many times the mean score difference of each sample was greater than that of the shuffled matrices. For each sample a p-value was computed as 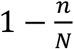 whereas *n* is the number of times the mean score difference of the particular sample was higher than that of the shuffled matrices and *N* is the number of repeats (1,000). The p-values were corrected for multiple testing for each program using the Bonferroni method.

### Assessment of the relative change in proportions of cell states in IDHi-treated vs. untreated samples

For each state *s* ∈ {*AC* − *like, OC* − *like, Undifferentiated/NPC* − *like, Cycling*}and for each pair of IDHi-treated and untreated samples, *t* ∈{*MGH170, MGH229*} and *u* ∈{*MGH36, MGH53, MGH54, MGH60, MGH93, MGH97*}, we defined *Ft, s* and *Fu, s* as the fraction of cells assigned to state *s* in samples *t* and *u* respectively and computed 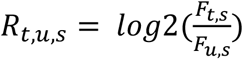 which represents the relative difference in the proportion of cells assigned to state *s* in the IDHi-treated sample *t* compared with the untreated sample *u* in log2FC units. Statistical significance of the difference was tested with a hypergeometric test (fisher.test function implemented is R’s stats package) for each *(s, t, u)* combination and corrected for multiple testing using the Benjamini-Hochberg method (p.adjust function implemented in R’s stats package).

### Processing of matched pre- and post-treatment samples for single-cell RNAseq

For each sample, a piece of frozen tumor tissue (approximately 5mm x 5mm x 5mm) was placed into a well of a 6-well plate (Corning, Cat. No. CLS3516-50EA) containing 1 ml of CST buffer and processed on ice. Tumor tissue was chopped for 10 minutes using Noyes Spring Scissors (Fine Science Tools, Cat. No. 15514-12). The sample was then filtered through a 40 um cell strainer (Fisher Scientific, Cat. No. 22363547) and the well and filter were washed with an additional 1 ml of CST buffer. The total volume was then brought up to 5 ml with 3 ml of ST buffer and the sample was centrifuged in a 15 ml conical tube at 4°C for 5 minutes at 500 g. The supernatant was removed and the pellet was resuspended in 100 ul ST buffer. The sample was then filtered through a 35 um cell strainer (Falcon, Cat. No. 352235). Nuclei suspension was counted using disposable C-Chip Hemocytometers (INCYTO, Cat. No. DHCN012). Single nuclei were processed through the 10X Chromium 3’ Single Cell Platform using the Chromium Single Cell 3’ Library, Gel Bead and Chip Kits, following the manufacturers protocol. Briefly, 8,000 single nuclei were loaded into each channel of the chip to be partitioned into Gel Beads in Emulsion (GEMs) in the Chromium instrument, followed by nuclei lysis and barcoded reverse transcription of RNA in the droplets. This was followed by amplification, fragmentation, and addition of adaptor and sample index. Libraries from two 10x channels were pooled together and sequenced on one lane of an Illumina Next-Seq 500 sequencer with paired end reads, Read 1: 28 nt, Read 2: 55 nt, Index 1: 8 nt, Index 2: 0 nt. Buffers for nuclei preparation were prepared as follows. ST buffer: Salt-Tris solution containing 146 mM NaCl (Thermo Fisher Scientific, Cat. No. AM9759), 10 mM Tris-HCl pH 7.5 (Thermo Fisher Scientific, Cat. No. 15567027), 1 mM CaCl2 (SigmaAldrich, Cat. No, 21115) and 21 mM MgCl2 (Sigma Aldrich, Cat. No. M1028) in ultrapure water. CST buffer: 10 ml of ST buffer, 320 µl of 0.25 M CHAPS (Thermo scientific, Cat. No. 28300) and 10 µl of 10% BSA (Sigma Aldrich, Cat. No. A3059).

### Single-cell RNAseq data processing of matched pre- and post-treatment samples

Illumina sequencing base calls were converted into FASTQ files using *cellranger* v4.0.0 pipeline (*mkfastq* command). Gene expression matrices were generated using *cellranger* (*count* command) by aligning the FASTQ files to a pre-mRNA reference transcriptome (GRCh38 GENCODE v32/Ensemble 98) which was built according to instructions provided by 10x Genomics. Each of the matched pairs was analyzed independently (pre- and post-treatment samples from the same pair were analyzed together) and the same analysis steps were applied to both of them. Expression data of each matched pair was normalized, clustered and annotated (for cell types) using the *Seurat* package(49). Malignant cells were detected using CNA analysis and classified into cell states as described above.

### Processing of in-vitro models for single-cell RNAseq

Patient derived primary IDH mutant glioma cultures (MGG119, MGG152, BT142) were grown in Neurobasal Medium (GIBCO 21103-049) supplemented with 1X N2/B27 (GIBCO), 1% Penicillin/Streptomycin (GIBCO), 1X Glutamax (GIBCO), 20 ng/mL EGF and 20 ng/mL bFGF (FGF2). The details of our cellular models have been published previously(13,29,38). IDHi treatment was performed by addition of 5 μM AGI-5198 (Cayman Chemical) to medium for at least 5 weeks. Treated and untreated samples were barcoded through TotalSeq™ DNA-tagged Cell Hashing antibodies (BioLegend) and multiplexed prior to processing through the 10X Chromium 3’ Single Cell Platform, following manufacturer’s instructions. Briefly, ∼1,000,000 cells per sample were resuspended in cold Cell Staining Buffer (BioLegend) containing 0.035% dextran sulfate and blocked for 10 min at 4 °C with TruStain FcX (BioLegend). Cell suspensions were subsequently stained with barcoded antibodies (TotalSeq™–B antibodies, BioLegend) for 30 min at 4 °C. Cells were washed with cold Cell Staining Buffer (BioLegend), resuspended in PBS containing 0.04% BSA and filtered through a 40 μm cell strainer. Four to six samples were pooled at equal concentration and processed in parallel through the 10X Chromium 3’ v3.1 Single Cell Platform using the Chromium Single Cell 3’ Library, Gel Bead and Chip Kits, following the manufacturers protocol. Briefly, ∼8,000 single cells were loaded into each channel of the chip to be partitioned into Gel Beads in Emulsion (GEMs) in the Chromium instrument, followed by cell lysis, barcoded reverse transcription of RNA in the droplets, and cDNA amplification for ten cycles. cDNAs derived from cellular mRNA were size separated from antibody-derived tags (ADTs) with SPRIselect Beads (Beckman Coulter, USA). Gene expression libraries were constructed by fragmentation of cDNAs derived from cellular mRNA and addition of adaptor and sample index. ADTs were amplified for ten additional cycles and ADT libraries were constructed by addition of sample index. ADT and cDNA libraries were pooled followed by Illumina sequencing.

### Single-cell RNAseq data processing of in-vitro models

Gene expression matrices were generated from Illumina sequencing base calls using the cellranger pipeline (mkfastq and count). Each of the models was analyzed independently of the other models (IDHi-treated and untreated cells from the same model were analyzed together) and the same analysis steps were applied for each of them. UMI counts for each cell were converted to *counts per million* units by dividing the gene counts of each cell by the “per million” scaling factor (total counts per cell divided by 1,000,000) and then log2-transformed. Cells were assigned to the respective sample-of-origin and treatment condition by taking the maximal value for that cell in the hashing table outputted by cellranger. Cells were considered as unreliably assigned if the difference between the maximal entry and the second best entry was less than two folds, in which case they were discarded from downstream analysis. Cells were excluded based on two additional quality control metrics: minimal complexity cutoff of 1000 genes and expression of ribosomal and mitochondrial genes (**Table S4**). Genes were filtered out based on minimal average expression cutoff of 4 as described above for patient samples. Next, cells were scored for the three cellular state programs and for the cell cycle programs. Cells were assigned to a state if the maximal score was greater than zero and the difference between the maximal score and the second best score was at least 0.1. To exclude the possibility that the relative differences between IDHi-treated and untreated cells in the fractions of cells assigned to a particular state are threshold dependent we conducted a sensitivity analysis (**Fig. S3E, Table S4**) by computing these relative differences for multiple potential threshods in the range [0, 0.3]. The fraction of cells defined as differentiated to either the AC-like or OC-like states varies extensively across this range, while the increased fraction of differentiated cells, in IDHi-treated vs. untreated samples, remained stable across the entire range (**Fig. S3F, Table S4**). Thresholds for G1/S and G2/M classification were determined by computing the sampling distribution of mean G1/S and G2/M scores and taking twice the standard error, which converged to ∼0.5 for all models. Sampling distribution of the mean G1/S and G2/M scores were computed by generating 100 shuffled gene expression matrices (by shuffling the value of each gene), and computing the mean G1/S and G2/M scores for each shuffled matrix.

### TCGA data preprocessing

Gene expression data (raw counts), clinical annotation file and mutation annotation file for the TCGA-LGG project were downloaded using the R package *TCGAbiolinks*(50). We filtered out genes with low variability, leaving 10,000 genes for analysis. Raw gene counts were transformed into counts-per-million (CPM) scale and log2-transformed as log2(CPM+1). We filtered out samples that do not have the 1p/19q co-deletion based on the clinical annotation file, leaving us with 150 samples. Since tumor grade may impact certain aspects of the analysis we filtered out 16/150 samples from the dataset for which grade annotation was not available in the clinical annotation file. Finally, we defined relative expression levels for each dataset separately by gene-wise centering of the *log2(CPM+1)* expression values.

### Bulk scoring of TCGA samples

We scored the TCGA samples using the centered log2(CPM+1) values for the AC-like, OC-like, NPC-like and cell-cycle programs (same gene-sets that were used for scoring the single-cell data) by computing the mean expression per sample for each program.

### TCGA AC differentiation analysis

We computed for each sample an AC-differentiation score, defined as 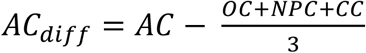 where *AC, OC, NPC* and *CC* represent the sample’s scores for the AC-like, OC-like, NPC-like and cell-cycle programs. This enabled deriving an expected AC differentiation score for each tumor grade *g* ∈{*Grade* 2,*Grade* 3} which is the average AC differentiation score across all samples that belong to grade *g*. Since tumor grade may impact the degree of AC-like differentiation, we compared the differentiation score distributions of grade 2 and grade 3 tumors and indeed found that grade 2 tumors were, on average, more differentiated than grade 3 tumors (p=0.0008, Wilcoxon rank sum test). We defined bulk expression profiles for each of the samples analyzed by scRNAseq by averaging across the cells in each tumor and computed AC differentiation scores for these samples as well using the bulk expression profiles. These scores span the expected spectrum of AC differentiation within the TCGA cohort (**Fig. S5A**). This property enabled using the expected AC differentiation computed from the TCGA to compute relative AC differentiation scores and account for differences in tumor grade. These scores were defined for each sample *i* as *ACdiff*_*i*_ *- E(ACdiff)* where *E(ACdiff)* is the expected AC differentiation score corresponding to sample *i*’s tumor grade (**Fig. S5B**).

### Mutations associated with changes in AC differentiation

TCGA data was separated into two datasets according to tumor grade. Each dataset was row-centered, scored for the AC-like, OC-like and NPC-like programs and the AC differentiation score was computed. We included in this analysis only mutations with at least 5 occurrences in a particular dataset (according to the mutation annotation file). For each mutation we computed the difference between the mean AC differentiation scores of the samples with the mutation and those without the mutation (relative AC differentiation score) and the statistical significance of the difference between the two groups using Wilcoxon rank sum test (corrected for multiple testing using the Benjamini-Hochberg method).

## Data Availability

All data for untreated patients produced in the present study are available upon reasonable request to the authors. All data for IDHi-treated patients produced in the present study will be made available at a later time.

## Acknowledgements

This work was supported by a Broad Institute–Israel Science Foundation Collaborative Project Award (I.T. and M.L.S.), the MGH Research Scholars Award (M.L.S.), grants from the Mark Foundation Emerging Leader Award (M.L.S.), the Sontag Foundation Distinguished Scientist Award (M.L.S.), N.I.H. R37CA245523 (M.L.S) and R01CA258763 (M.L.S.). I.T. is the incumbent of the Dr. Celia Zwillenberg-Fridman and Dr. Lutz Zwillenberg Career Development Chair, and is supported by the Zuckerman STEM Leadership Program, the Mexican Friends New Generation, the Benoziyo Endowment Fund, and grants from the Human Frontiers Science Program. A.S. is partially supported by the Israeli Council for Higher Education (CHE) via the Weizmann Data Science Research Center, and by a research grant from the Estate of Tully and Michele Plesser.

## Author contributions

A.S., S.G., D.P.C., M.L.S. and I.T. conceived the project, designed the study, and interpreted results. S.G. and H.R.W. collected oligodendroglioma single cells and generated sequencing data. A.S., R.C. and I.T. performed computational analyses. J.M.F., R.L.S., and R.M. provided flow-cytometry expertise. L.N.G.C., D.P.C., J.J.M, I.A.R., P.Y.W., W.P., and K.L.L. identified and consented patients for the study. C.K.L., H.N., H.W. and D.N.L. provided experimental and analytical support. I.T., D.PC. and M.L.S. jointly supervised this work and interpreted results. A.S., S.G., D.P.C., M.L.S. and I.T. wrote the manuscript with feedback from all authors.

## Competing interests

M.L.S. is equity holders, scientific co-founder and advisory board member of Immunitas Therapeutics. I.T. is advisory board member of Immunitas Therapeutics. D.P.C. has consulted for Lilly and Boston Pharmaceuticals, and has received honoraria and travel reimbursement from Merck for invited lectures. The authors declare that such activities have no relationship to the present study.

## Notes

### Author Declarations

Institutional Review Board 10-417 of Dana-Farber Harvard Cancer Center gave ethical approval for this work

