## Supplemental Figures 1-7 for "Mutant IDH Inhibitors Induce Lineage Differentiation in IDH-mutant Oligodendroglioma"

Supplemental Figure 1

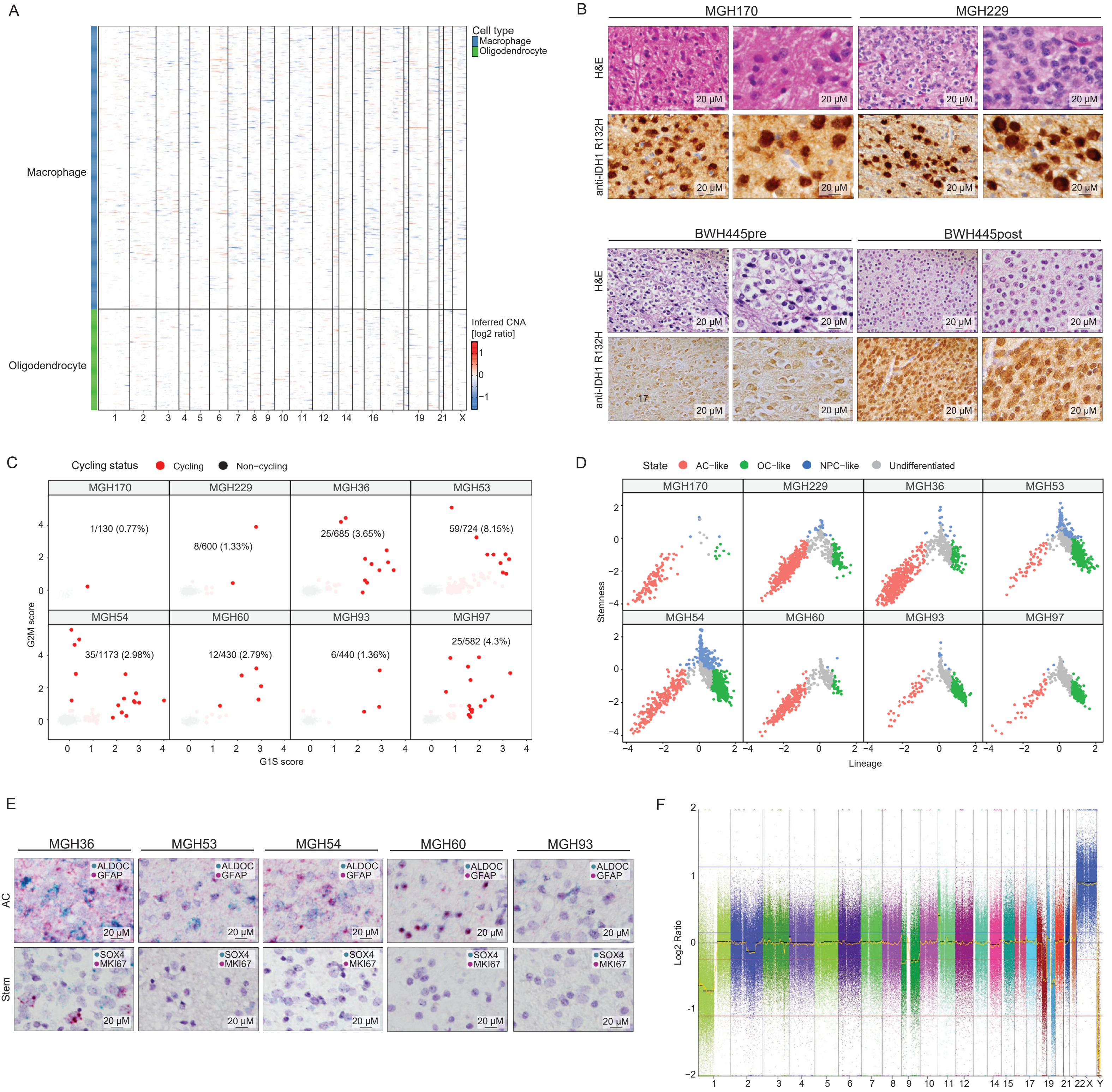

**Figure S1.**

(**A**) Copy-number aberrations inferred from single-cell expression data of cells classified as non-malignant (shown for unmatched cohort only). Rows represent cells and columns represent chromosomal locations. As expected, non-malignant cells lack 1p/19q-codel signal as well as other copy-number aberrations. (**B**) Representative H&E and anti-IDH1 R132H staining in the IDHi-treated samples (MGH170, MGH229 and BWH445). (**C**) Each panel shows the malignant cells from one tumor (unmatched cohort), scored for the G1/S (X axis) and G2/M (Y axis) cell cycle programs. Cells are colored by their classification as cycling (red) or non-cycling (black), and the fraction of cycling cells is indicated. (**D**) Each panel shows the malignant cells from one tumor (unmatched cohort), scored for stemness (Y axis) and lineage (X axis). Lineage score is defined as the maximum between the OC-like score and AC-like scores. Stemness score is defined as the difference between the NPC-like and the Lineage scores. Cells are colored by their assignment to four cellular states (see *Methods*). (**E**) *In situ* RNA hybridization of IDHi untreated samples for AC-like (*GFAP*, *ALDOC*), Stem-like (*SOX4*), and proliferation (Ki67) markers. (**F**) Array comparative genomic hybridization showing 1p/19q whole-arm co-deletion in tumor BWH445 (on-treatment).

### Supplemental Figure 2

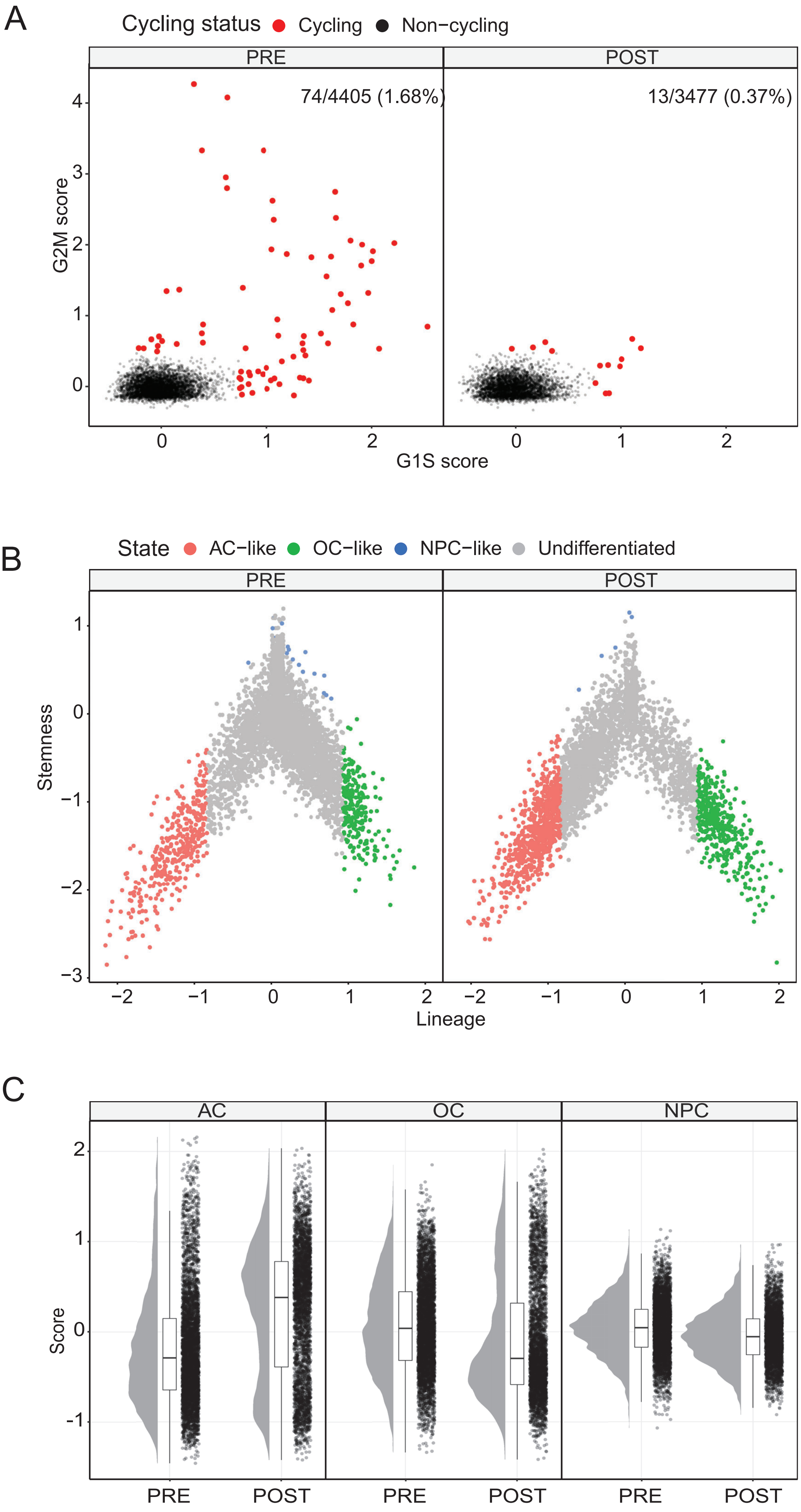

**Figure S2**

(**A**) Each panel shows the malignant cells from either on- or pre-treatment matched samples, scored for the G1/S (X axis) and G2/M (Y axis) cell cycle programs. Cells are colored by their classification as cycling (red) or non-cycling (black), and the fraction of cycling cells is indicated. (**B**) Each panel shows the malignant cells from either on- or pre-treatment matched samples, scored for stemness (Y axis) and lineage (X axis). Lineage score is defined as the maximum between the OC-like score and AC-like scores. Stemness score is defined as the difference between the NPC-like and the Lineage scores. Cells are colored by their assignment to four cellular states (see *Methods*). (**C**) Cell state score distribution for either on- or pre-treatment matched samples. Each dot represents a cell.

### Supplemental Figure 3

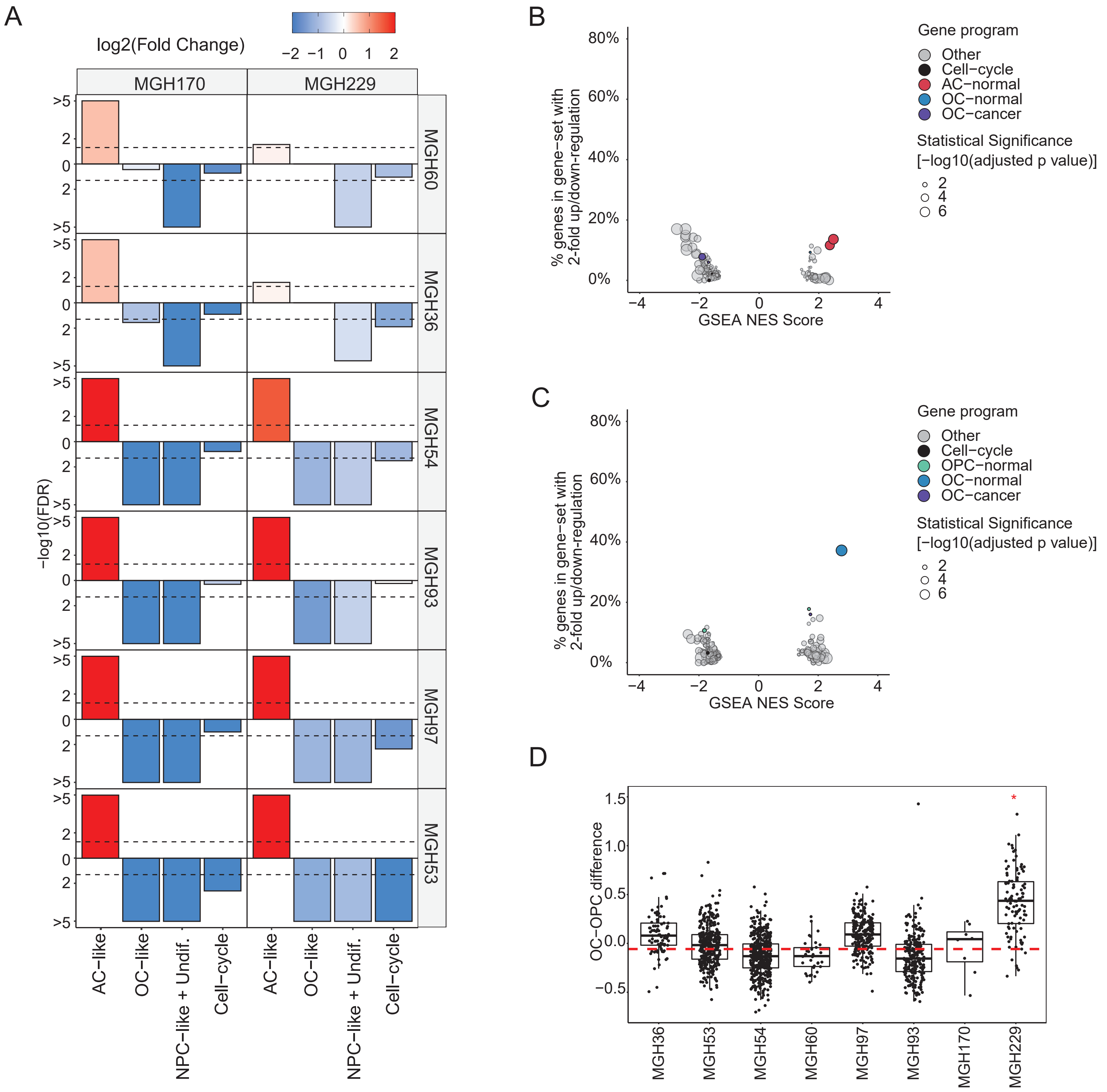

**Figure S3**

(**A**) Pairwise comparisons between each of the IDHi-treated and untreated samples (unmatched cohort) across the different cellular states. Bar values represent the statistical significance of each pairwise comparison, defined as -log10 of a p-value calculated by hypergeometric test and corrected for multiple testing using the Benjamini-Hochberg method; bar direction (up or down) is defined by an increase or decrease, respectively, of the relevant state in the IDHi-treated sample Bar colors represent the relative change in state fractions (computed as $log2(\frac{\text{Fraction in treated}}{\text{Fraction in untreated}})$. This figure demonstrates a statistically significant decrease in the fraction classified either as Undifferentiated or NPC-like in all 12 pairwise comparisons as well as a statistically significant increase in the fraction of AC-like cells in 10/12 pairwise comparisons. (**B**) Comparison of IDHi-treated and untreated samples (unmatched cohort) limited only to the cells classified as AC-like in each sample using GSEA. Each dot represents a gene-set that passed the statistical significance threshold (adjusted p-value < 0.05), dot size represents the extent of statistical significance and dot color indicates whether the gene-set belongs to any of the glioma hierarchy/neural development gene-sets. X-axis shows the GSEA Normalized Enrichment Score (NES), Y-axis shows the fraction of genes in each gene-set with an absolute log2-ratio greater than 1 (e.g. genes in the extreme ends of the ranked list used for GSEA computation with more than a twofold change). (**C**) Same as panel B but here with samples limited to cells classified as OC-like. (**D**) Cells were placed on an OPC-normal to OC-normal differentiation axis by computing the difference between the two scores and the distribution of score differences is shown. Cells classified as OC-like that belong to MGH229 are significantly more differentiated towards the OC-normal state ($p<10^{-3}$, permutation test, see *Methods*) than cells that belong to the untreated samples. Red horizontal line indicates the median score of all untreated samples, red asterisk indicates statistically significant samples.

### Supplemental Figure 4

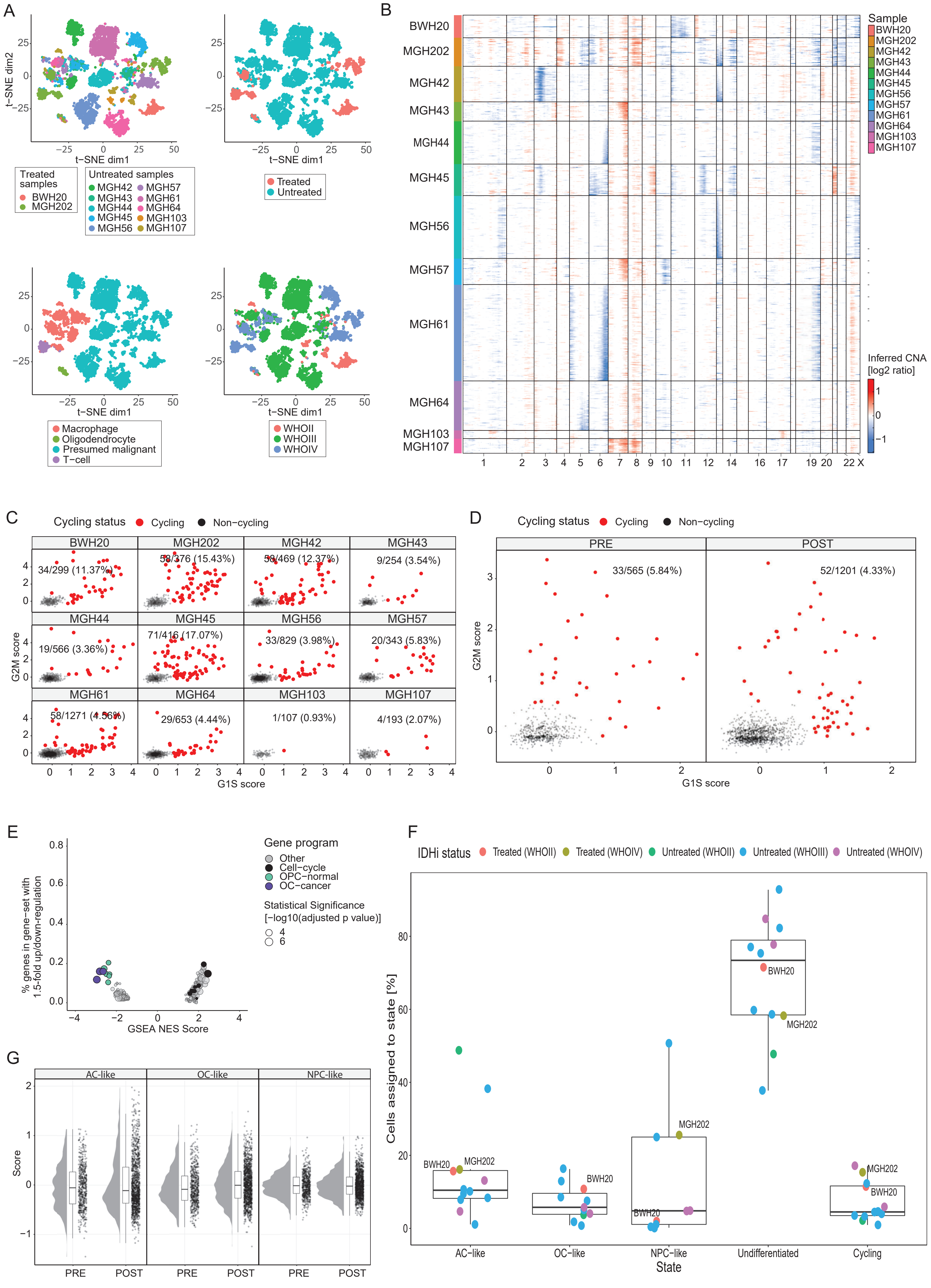

**Figure S4 - The non-responder cohort**

(**A**) t-distributed stochastic neighbor embedding (t-SNE) plots showing 5,487 single cell expression profiles of the unmatched *non-responder* cohort. Each of the four plots shows the same cells and coordinates, but colored in a different way as labeled below the figure, with colors corresponding to (from left to right): sample identity, treatment status, cell type and tumor grade. (**B**) Copy-number aberrations inferred from single-cell expression data (unmatched cohort) of cells classified as malignant. Rows represent cells and columns represent chromosomal location. (**C**) Each panel shows the malignant cells from one tumor (unmatched cohort), scored for the G1/S (X axis) and G2/M (Y axis) cell cycle programs. Cells are colored by their classification as cycling (red) or non-cycling (black), and the fraction of cycling cells is indicated. (**D**) Same as panel C but here showing the matched pre- and post-treatment samples of BWH5033. (**E**) Comparison of IDHi-treated and untreated samples (unmatched cohort) using GSEA. Each dot represents a gene-set that passed the statistical significance threshold (adjusted p-value < 0.05), dot size represents the extent of statistical significance and dot color indicates whether the gene-set belongs to any of the glioma hierarchy/neural development gene-sets. X-axis shows the GSEA Normalized Enrichment Score (NES), Y-axis shows the fraction of genes in each gene-set with an absolute log2-ratio greater than 1 (e.g. genes in the extreme ends of the ranked list used for GSEA computation with more than a twofold change). (**F**) Percent cells assigned to each state in each sample (unmatched samples). Each dot represents a sample and colored according to tumor grade and treatment status. There is no statistically significant difference between the treated and untreated samples in terms of percent cells assigned to any of the states (p=0.27, 0.54, 0.66, 0.6 and 0.18 for the AC-like, OC-like, NPC-like, Undifferentiated and Cycling states, respectively, Wilcoxon’s rank sum test). (**G**) Cell state score distribution for the matched pre- and post-treatment samples of BWH5033. Each dot represents a cell.

### Supplemental Figure 5

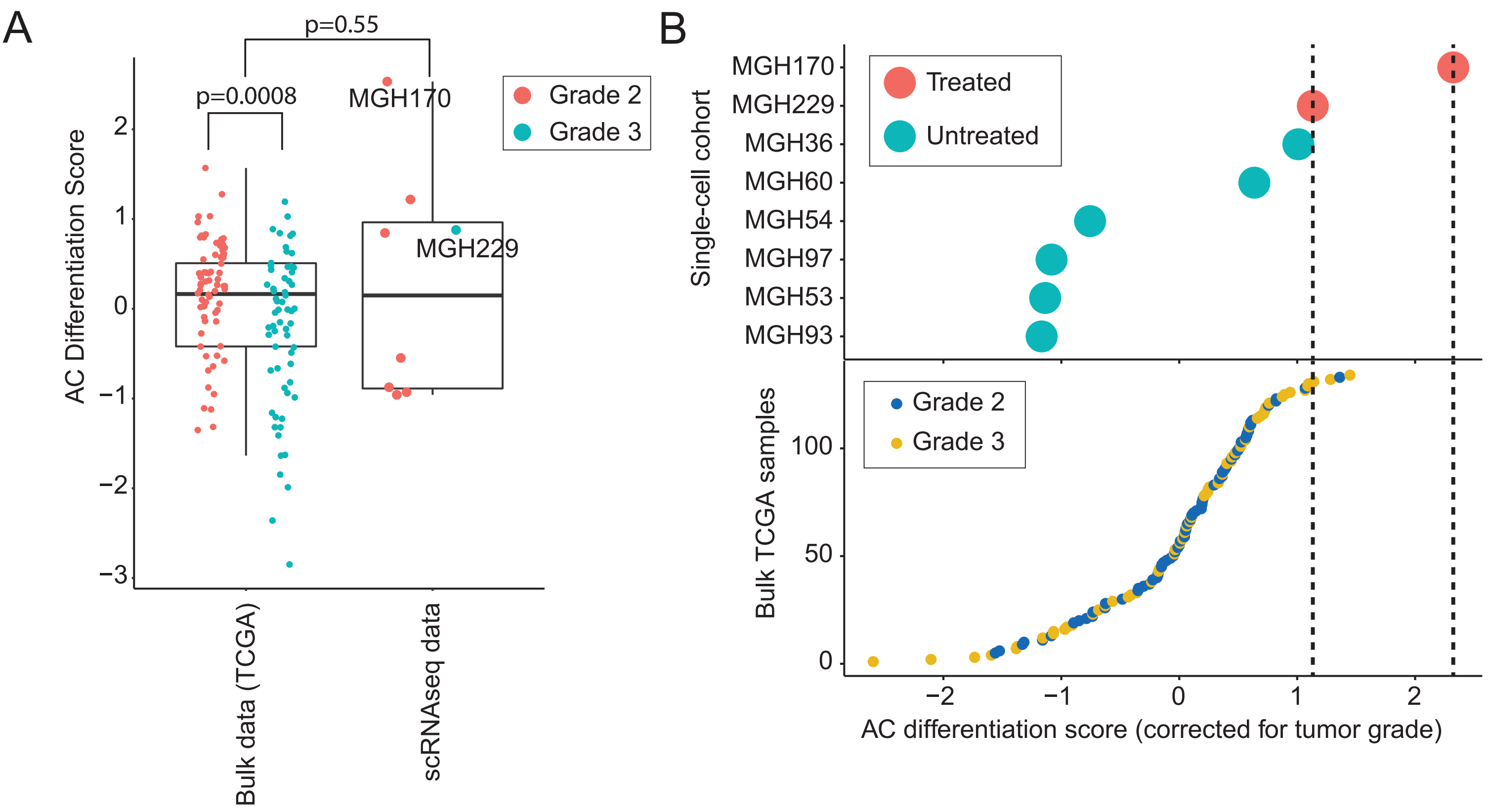

**Figure S5 - TCGA analysis**

(**A**) The distribution of AC differentiation scores for 8 scRNAseq and 134 TCGA samples stratified by tumor grades (72 grade 2, 62 grade 3). The aggregate AC differentiation scores of the scRNAseq samples were computed by averaging across all cells in each tumor. These largely span the spectrum of TCGA AC differentiation scores (p=0.55 for difference between TCGA and scRNAseq score distributions, two-sided Wilcoxon's rank sum test), supporting the use of the expected AC differentiation derived from the TCGA scores distribution as a measure for score normalization. AC differentiation scores are higher in grade 2 samples compared to grade 3 samples (p=0.0008, Wilcoxon's rank sum test, one sided). (**B**) AC differentiation scores corrected for tumor grade (see Methods) in the single cell cohort (top panel) and in the bulk oligodendroglioma cohort from TCGA (bottom panel). None of the TCGA samples score as highly as MGH170 and only 3% (4/134) score as highly as MGH229.

Supplemental Figure 6

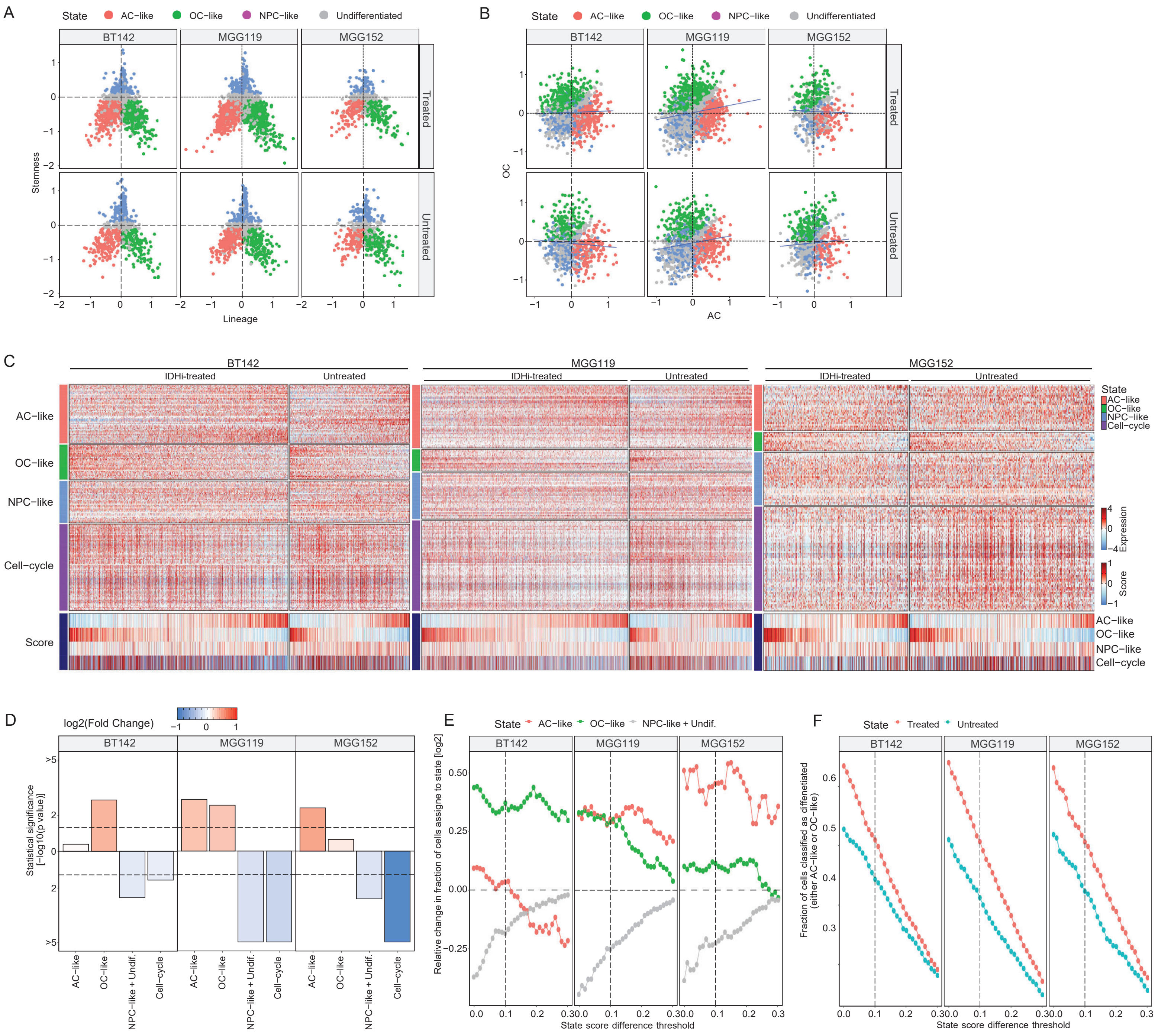

**Figure S6 - Validation using models (IDH mutant gliomaspheres, non 1p/19q co-deleted)**

(**A-B**) Each panel shows cells from one *in-vitro* model (as indicated by column labels) either before or after IDHi treatment (as indicated by row label), with cells scored for lineage and stemness (in A) or for the AC-like and OC-like programs (in B). (**C**) Heatmap showing the expression of genes associated with the AC-like, OC-like and NPC-like programs, after excluding lowly expressed genes in each model. Cells are ordered by AC-like score minus OC-like score and grouped by IDHi treatment status; genes are ordered by hierarchical clustering. Bottom panel shows the scores for the three cellular states and for cell-cycle. (**D**) Each panel shows in one *in vitro* model, changes in the fraction of cells assigned to each cellular state, between treated and untreated samples. The presentation is similar to that in **Fig. S2A**: Bar values represent statistical significance defined as -log10 of a p-value calculated by hypergeometric test and corrected for multiple testing using the Benjamini-Hochberg method; bar direction (up or down) is defined by an increase or decrease, respectively, of the relevant state in the IDHi-treated sample. Bar colors represent the relative change in state fractions (computed as $log2(\frac{\text{Fraction in treated}}{\text{Fraction in untreated}})$. The figure shows a small but consistent and statistically significant decrease in the fractions of NPC-like, undifferentiated and cycling cells in the IDHi-treated samples. The IDHi effect on the differentiated cells is less consistent but in each of the models at least one lineage is enriched in the IDHi-treated sample. (**E**) Sensitivity analysis of the state classification algorithm. X-axis shows potential values for the difference in cell scores that is used as a threshold for assignment of cell states. Y-axis shows the relative change in assignment of cells to a certain state between IDHi-treated and untreated samples (same as the color-coded values in panel D), when using the threshold indicated on the X-axis; this analysis shows highly consistent results (i.e. most values are consistently below or above zero across all thresholds) demonstrating that the results described in panel D are threshold-independent.

Supplemental Figure 7

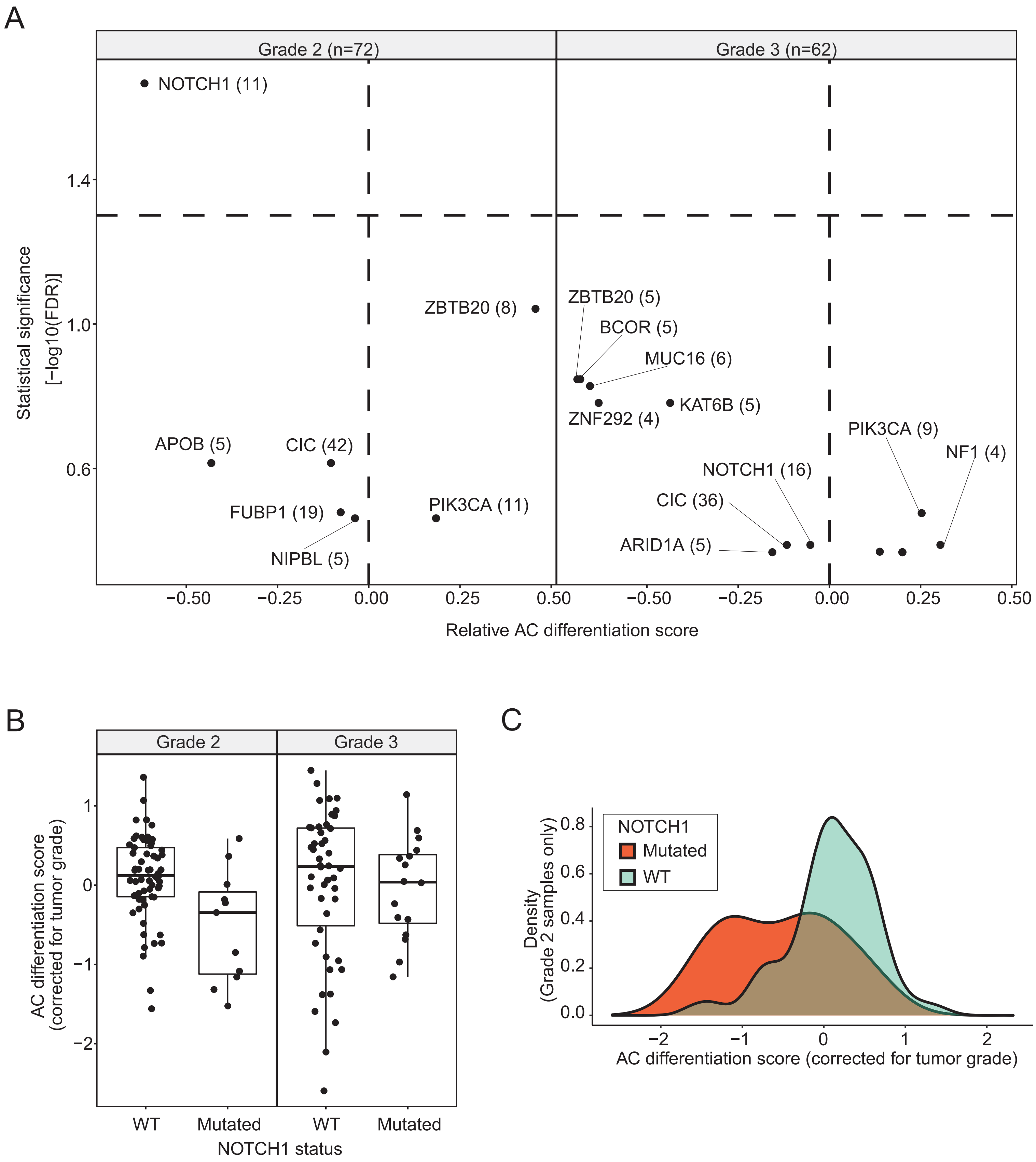

**Figure S7 - NOTCH1 mutations are associated with decreased AC differentiation scores in grade 2 samples**

(**A**) Analysis of the association between recurrent mutations (at least in five tumors) and AC differentiation scores, for oligodendroglioma tumors of grade 2 (left) and of grade 3 (right) reveals a significant association (FDR-adjusted p-value=0.02, Wilcoxon rank sum test) between mutations in *NOTCH1* and low degree of AC differentiation in grade 2 lesions (11/72 samples) but not in grade 3 lesions (16/62 samples). Each panel shows the difference in average AC differentiation score between tumors with and those without a specific mutation (X-axis), and the significance of that score (Y-axis, defined by -log10 of the p-value calculated by t-test and corrected for multiple testing by the Benjamini-Hochberg method). Horizontal line shows a significance threshold (FDR=0.05), highlighting NOTCH1 in grade 2 oligodendroglioma as the only significant association; Vertical line represents the mean AC differentiation score. (**B**) Distribution of the AC differentiation scores (corrected for tumor grade) shows a significant difference in the scores of grade 2 *NOTCH1* mutants. This effect is not seen in grade 3 samples. (**C**) Density of AC differentiation scores (corrected as in (**Fig. S5B**)) among grade 2 oligodendroglioma samples with wild-type (green) or mutant (orange) *NOTCH1*.
